# Shining a Light on Athletes’ Sleep: Development of a Screening Nomogram to Flag Athletes at Risk of Poor Sleep Quality

**DOI:** 10.64898/2026.03.04.26347647

**Authors:** Shauna Stevenson, Matthew Driller, Hugh Fullagar, Kate Pumpa, Haresh Suppiah

**Affiliations:** Sport, Performance, and Nutrition Research Group, School of Allied Health, Human Services, and Sport, La Trobe University, Melbourne, Australia; Physical Activity, Physical Education, Sport and Health (PAPESH) Research Centre, Sports Science Department, Reykjavik University, Reykjavik, Iceland; Aspetar Orthopaedic and Sports Medicine Hospital, Doha, Qatar; Institute for Sport and Health, University College, Dublin, Ireland

**Keywords:** Circadian rhythms, light exposure, sport, recovery, LASSO regression

## Abstract

**Background:** Emerging research indicates that light exposure may influence sleep quality. Identifying key light-exposure behaviours associated with poor sleep quality in athletes may allow practitioners to efficiently screen for sleep difficulties and prioritise athletes for further assessment. Translating these findings into a practical screening tool could enhance willingness of high-performance professionals to monitor sleep and light exposure in athletes.

**Hypothesis:** Key predictor variables identified by feature reduction techniques will lead to higher predictive accuracy in determining which light behaviours are associated with poor sleep quality in athletes.

**Study Design:** Cross-sectional study.

**Level of Evidence:** Level 3.

**Methods:** 121 athletes from varying competitive levels completed questionnaires, including the Light Exposure Behaviour Assessment (LEBA) and Pittsburgh Sleep Quality Index (PSQI). Poor sleep quality was defined using the PSQI cut-off >5. Least absolute shrinkage and selection operator (LASSO) regression identified light exposure variables from the LEBA questionnaire most strongly associated with good and poor sleep quality in athletes. Three models were compared: a full-variable model (23 items), a factor-specific model (Factor 3: screen/device use), and a feature-reduced model (LASSO-selected items).

**Results:** Phone use before bed, checking phone/watch during the night, were identified as variables of greatest association with poor sleep quality and used for reduced feature set modelling. On an independent test set, the feature-reduced model achieved area under the curve (AUC) 0.83, sensitivity 0.70, and specificity 0.92.

**Conclusions:** Our findings report that phone-related behaviours before and in bed are associated with a higher likelihood of poor sleep quality. These behaviours, combined with the developed nomogram, provide a preliminary, low-burden screening tool to identify athletes who may be experiencing sleep difficulties. The high specificity indicates that athletes flagged by the tool are likely to have genuine poor sleep quality, warranting further assessment to identify underlying causes and appropriate interventions.

**Clinical Relevance:** Education and interventions focused on light exposure factors were identified as most influencing sleep quality in a multifaceted athletic population and could be prioritised to optimise sleep quality. The developed sleep quality nomogram may be useful as a decision-making tool to improve sleep monitoring practice among practitioners.

## Introduction

Elite athletes frequently fail to achieve the recommended amount of sleep per night (Watson, 2017) with athletes from various sports demonstrating differing rest-activity circadian rhythms (Vitale et al., 2019). One of the primary cues regulating circadian rhythms is light exposure (Duffy & Czeisler, 2009). An athlete’s light-dark cycle may be disrupted by factors such as irregular training and competition schedules (Sargent et al., 2014), and varying training environments (e.g., indoor vs outdoor settings). Previous research has utilised objective light exposure measures to examine associations with sleep outcomes (Didikoglu, et al., 2023; Scheuermaier et al., 2010; Stevenson et al., 2024b), and has consistently highlighted a potential link between light exposure and sleep quality. However, such measures present practical limitations, including large data processing requirements, time-consuming analysis, and may be difficult to implement in large athletic populations. Therefore, a more robust, practical tool that best discriminates light-related behaviours most relevant to sleep outcomes is still required, particularly in applied high-performance sport settings.

Light exposure is a key environmental cue that entrains the human circadian system (Corbett et al., 2012), which regulates fundamental physiological processes, including sleep and core body temperature (Hastings et al., 2008). The human circadian system integrates variations in light exposure throughout the day with increased morning and daytime light, and reduced evening light, supporting healthier sleep patterns (Duffy & Wright, 2005; Knufinke et al., 2019a). Insufficient sleep quality in athletes arises from a combination of both sport- and non-sport-related factors (Walsh et al., 2021a), including suboptimal light exposure (Figueiro et al., 2017). A recent study conducted in elite athletes reported that higher levels of morning light exposure were associated with increased total sleep time, and increased daytime light exposure was significantly correlated with better subjective sleep quality (both *p* < 0.05) (Stevenson et al., 2024). Further, increased evening light exposure in the lead-up to sleep has been linked to greater sleep disturbances (Stevenson et al., 2024). Although restricting evening light exposure is commonly recommended within sleep hygiene practices (O’Donnell & Driller, 2017), such recommendations warrant careful evaluation before routine implementation (Jones et al., 2019) due to inconsistencies in the literature regarding whether sleep impairments are driven primarily by the spectral and intensities of light exposure or by the cognitive engagement of evening activities (e.g., smartphone use) (Hartstein et al., 2024; Vézina-Im et al., 2025). Moreover, substantial inter-individual variability in light sensitivity and perception may further modulate these relationships (Chellappa, 2021). Other light-based interventions have shown encouraging results in recreational athletes. For instance, evening light restriction using amber-lens glasses improved sleep latency, sleep quality, and morning alertness in a group of recreational athletes (Knufinke et al., 2019b). Similarly, combined morning bright-light exposure and evening light restriction shortened subjective sleep latency by 8 minutes in recreational athletes, and by 17 minutes in those with rigid sleep-wake cycles (Knufinke et al., 2021).

While numerous studies have evaluated how light exposure interventions influence sleep outcomes, there remains a limited understanding of which specific light-related behaviours are most strongly associated with sleep quality in athletes, leading to a lack of practical tools that can be utilised in applied settings. Objective measures, such as wearable devices (often clip-on light sensors attached to clothing), can quantify light exposure (Scheuermaier et al., 2010; Stevenson et al., 2025; Stone et al., 2020), but are often impractical in sports environments due to logistical demands, sensor placement issues, clothing obstruction, and device errors. Self-report tools help mitigate these challenges but may include items that lack direct applicability to sleep-focused outcomes and limited relevance to athlete populations. For instance, athletes may experience substantial variability in light exposure within training or competition settings that combine outdoor and indoor components, meaning that broad time-based questionnaire items may lack sufficient precision in athletic populations and may also lack direct applicability to sleep-focused outcomes. For example, the Harvard Light Exposure Assessment (H-LEA) (Bajaj et al., 2011), provides a detailed resolution of timed light exposure throughout the day but relies on a comprehensive classification of light sources e.g., halogen versus fluorescent light, which may not be intuitive for athletes, thereby reducing practicality in athletic contexts. Additionally, the Light Exposure Behaviour Assessment (LEBA) (Siraji et al., 2022) provide insight into light-related habits and have demonstrated value for guiding light-based health interventions and predicting sleep quality (Siraji et al., 2023). However, this questionnaire has not yet been employed in athletic contexts, where compliance and behavioural specificity are key considerations. Feature-reduction methods may therefore be advantageous for identifying the most prominent light exposure items that meaningfully discriminate sleep quality, enabling practitioners to administer a rapid, targeted screen during consultations while maintaining predictive utility. In addition, visual decision-support tools such as nomograms, graphical calculators that translate predictor values into probability estimates, may help practitioners e.g., sport scientists, performance staff, or clinicians, interpret predictive models, identify athletes who are potentially more susceptible to poor sleep quality. Importantly, these tools can be applied with minimal time burden (typically <1-2 minutes) and facilitate the translation of research findings into practical recommendations, as demonstrated in previous work examining sleep hygiene predictors in adolescent athletes (Suppiah et al., 2022).

Previous research has highlighted a potential association between light exposure and sleep outcomes in the athletic population. Further, while light-based interventions are increasingly utilised in athletic settings, there is no concise tool that identifies which light behaviours are most strongly linked with sleep outcomes in athletes. While maintaining consistent sleep-wake rhythms and regulating daily light exposure has been recommended for athletic recovery and performance (Knufinke et al., 2021; Yin et al., 2021), the specific light behaviours that meaningfully relate to sleep quality in athletes remain unclear. Therefore, the present study aimed to identify which light-related behaviours from the LEBA questionnaire are most strongly associated with poor sleep quality in athletes, using LASSO regression for feature reduction. Using the most accurate predictive model, a secondary aim was to develop a nomogram as a practitioner-friendly visual tool to support the screening and interpretation of light exposure behaviours in the athletic population.

## Methods

In a cross-sectional study design, a total of 178 survey responses were assessed for eligibility. Of the 178 responses, 25 were excluded at screening due to sleep medication use, sleep disorders, supplements or medications affecting light perception. A further 32 responses were excluded prior to analysis due to missing questionnaire items or data quality issues, resulting in a final sample of 121 participants. Participants (50 female and 71 male; 28 ± 9 y) were from a multi-faceted athletic population (28 recreationally active, 30 trained/developmental, 40 highly trained/national level, 22 elite/international level or higher (McKay et al., 2022)). Participants were recruited from an array of different sports including Australian Football League (AFL) (n = 27), triathlon (n = 22), resistance training (n = 12), running (n = 9), Gaelic Athletic Association (GAA) (n = 5), soccer (n = 5), basketball (n = 4), swimming (n = 3), netball (n = 3), cricket (n = 2), cycling (n = 2), canoeing (n = 2), sailing (n = 2), lawn bowls (n = 2), unknown sport (n = 4). Athletes completed the survey across different season phases (29 in-season, 26 off-season, 43 pre-season, and 23 unspecified). Participants were from across 20 countries (Australia, Brazil, Colombia, Finland, France, Germany, Ireland, Italy, Mexico, New Zealand, Philippines, Portugal, Singapore, South Africa, Sweden, Switzerland, Taiwan, United Kingdom, United States and Vietnam). Participants were recruited via a combination of social media channels, word of mouth, and by targeting specific sporting organisations and national bodies (via email). The inclusion criteria for the current study were that they were free from pathological sleep disorders, visual impairments, or not taking any medication/supplements that may impact response to light, e.g., antidepressants or had any health complications. The study was approved by the institution’s Human Research Ethics Committee (HEC24435).

### Study Protocol

Once athletes met the inclusion criteria and provided informed consent via the survey URL link, access was granted to the survey. Participants were required to complete the survey via REDCap (Research Electronic Data Capture, Vanderbilt, USA) electronic data capture tools hosted at La Trobe University. A full description of how to complete each questionnaire was provided, and athletes were asked to complete all questionnaires simultaneously. The anonymised survey also included basic demographic information at the start (sleep medication or supplements, age, gender, sport type, competitive level and country).

### Survey Instruments

#### Light Exposure Behaviour Assessment (LEBA)

The Light Exposure Behaviour Assessment (LEBA) is a 23-item questionnaire containing five factors: F1, wearing blue light filters; F2, spending time outdoors, F3, using a phone and/or a smartwatch in bed; F4, using light before bedtime; and F5, using light in the morning and during daytime (Siraji et al., 2022). The questionnaire asks participants how frequently they engaged in specific behaviours over a timeframe of the past four weeks using a 5-point Likert scale (1 = never, 2 = rarely, 3 = sometimes, 4 = often, and 5 = always). Each factor is scored by adding up all corresponding item scores (Siraji et al., 2022). The LEBA questionnaire has been deemed successful in predicting chronotype, mood, and sleep quality in the general population (Siraji, Spitschan, et al., 2023b).

#### Pittsburgh Sleep Quality Index (PSQI)

The Pittsburgh Sleep Quality Index (PSQI) is a 19-item self-rated questionnaire tailored to assess sleep quality and sleep disturbances in its participants over 4 weeks (Buysse et al., 1989) with subscales including measures of sleep quality, sleep latency, sleep duration, sleep efficiency, sleep disturbances, sleep medication consumption habits, and daytime dysfunction (Buysse et al., 1989). The PSQI scores range from 0 to 21, whereby higher scores (>5) are indicative of poorer sleep quality and have a diagnostic sensitivity of 89.6 % and specificity of 86.5 % when distinguishing between “good” and “poor” sleepers (Buysse et al., 1989). This tool has been utilised in previous studies to determine athletes’ sleep history and previous sleep disturbances based on subscale scores previously mentioned (Halson, 2019).

### Statistical Analysis

Statistical analysis was conducted using R statistical software (R 4.4.2, R Foundation for Statistical Computing). Poor sleep quality was defined as a Pittsburgh Sleep Quality Index (PSQI) global score greater than 5 (Buysse et al., 1989).

#### Feature Reduction: Least Absolute Shrinkage and Selection Operator

To identify the smallest subset of LEBA items that best discriminate between good and poor sleep quality whilst maintaining practical interpretability, we utilised least absolute shrinkage and selection operator (LASSO) regression for feature reduction. LASSO regression addresses overfitting and model performance overestimation by placing a penalty on the sum of coefficients, shrinking coefficients with minor contributions towards zero, effectively removing them from the model. This produces a parsimonious and interpretable model (McNeish, 2015).

A total of 23 LEBA items across 5 factors were used as predictor variables. Items were standardised (*z*-score) to reduce the influence of magnitude differences on coefficient shrinkage. Model fit was performed using the *glmnet* package (Friedman et al., 2010) in R with 10-fold cross-validation. Alpha was set to 1 (pure LASSO penalty, as opposed to ridge regression or elastic net), and the optimal lambda was determined at the minimum cross-validation error.

#### Data Preprocessing

Participants with complete data on all LEBA items and PSQI global score were included in the analysis (N = 121). Model training and evaluation were performed using the *caret* package (Kuhn, 2015). A generalised linear model (logistic regression) was used to predict sleep quality (poor vs good). An 80:20 split was used for training and test sets by stratified random sampling (training N = 98; test N = 23).

Three models were trained and evaluated: (1) a full-feature model using all 23 LEBA items, (2) a factor-specific model using the 5 items from Factor 3 (using a phone and smartwatch in bed) as LASSO regression selected items from this factor, and (3) a feature-reduced model using items selected by LASSO regression.

#### Model Validation and Evaluation

This study employed 5-repeated 10-fold cross-validation on the training set. Cross-validation performance determined the best classification model. Predictive accuracy was evaluated on the held-out test set. Area under the curve (AUC) in receiver operating characteristic, sensitivity, and specificity were used to evaluate model performance. The sum of sensitivity and specificity (Youden method) was used for threshold selection.

#### Nomogram

The *rms* package (Harrell Jr, 2023) in R was used to construct the nomogram based on the feature-reduced model with the best predictive accuracy, providing a practical clinical tool for estimating the chance of poor sleep quality. The analysis workflow is presented in Figure 1.

**Figure 1:**
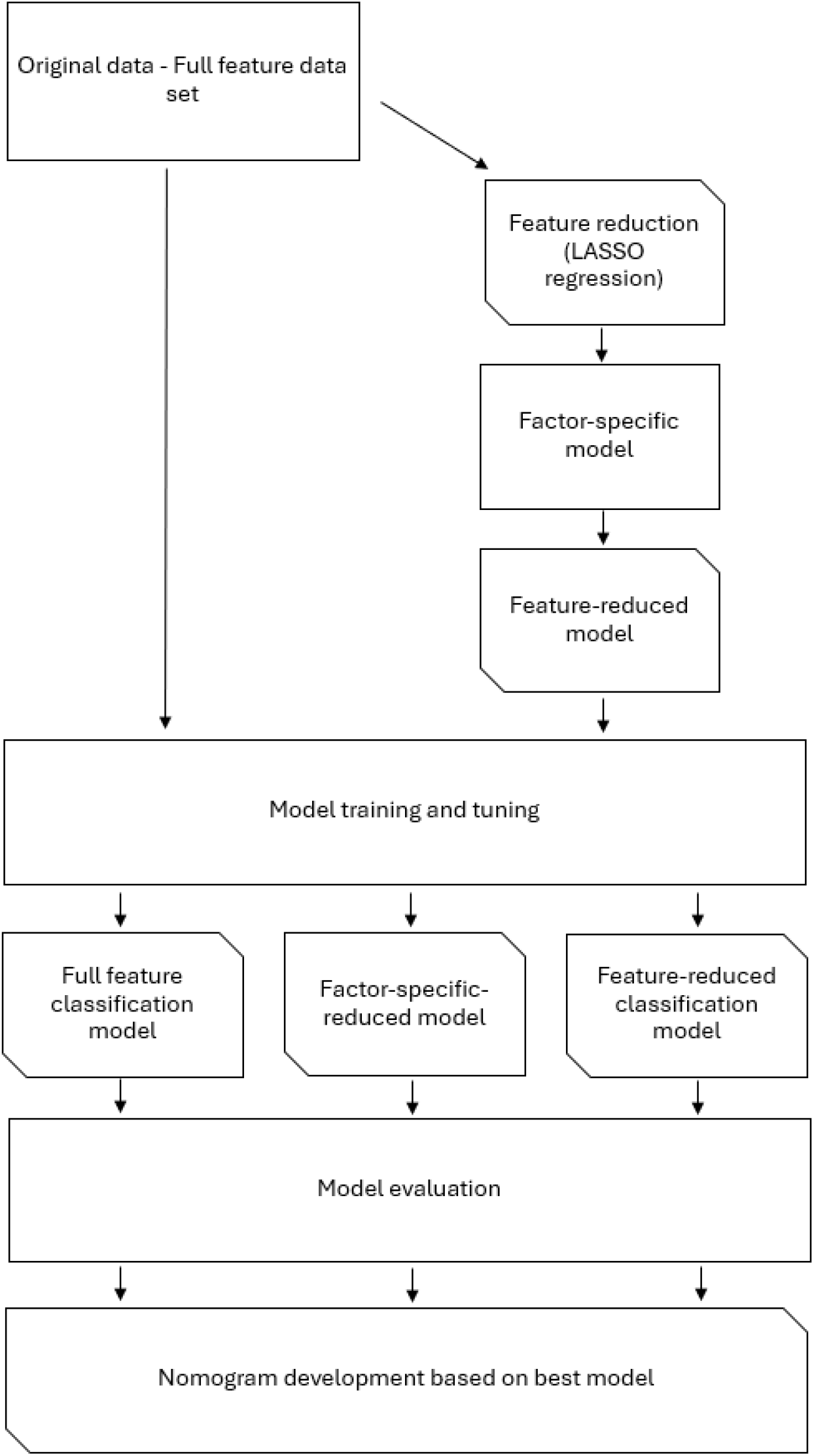
Feature selection and analysis workflow. LASSO, least absolute shrinkage and selection operator.

## Results

Descriptive statistics of LEBA factors and PSQI are displayed in Table 1.

**Table 1:** Descriptive statistics, including the mean and standard deviation of all LEBA factors and PSQI metrics of the participants in the current study.

| Variables | Mean ± SD |
| --- | --- |
| **Light Exposure Behaviour Assessment (LEBA):** |  |
| F1: Wearing blue light filters | 4.1 ± 2.3 |
| F2: Spending time outdoors | 19.6 ± 3.5 |
| F3: Using phone and smartwatch in bed | 15.4 ± 3.9 |
| F4: Using light before bedtime | 12.2 ± 4.3 |
| F5: Using light in the morning and during daytime | 10.7 ± 3.1 |
| **Pittsburgh Sleep Quality Index (PSQI):** |  |
| Bedtime (hh:mm) | 22:40 ± 1:12 |
| Waketime (hh:mm) | 6:42 ± 1:23 |
| Sleep Latency (min) | 23 ± 37 |
| Total Sleep Time (h:min) | 7 h 7 min ± 51 min |
| Sleep Efficiency (%) | 88.4 ± 8.4 |
| Sleep Disturbances (/5) | 1.0 ± 0.3 |
| Sleep Quality (/5) | 1.1 ± 0.6 |
| Sleep Medication (/5) | 0.1 ± 0.6 |
| Daytime Dysfunction (/5) | 1.1 ± 0.8 |
| Global Score (/21) | 5.5 ± 2.4 |

### Feature Reduction: LASSO

The LASSO regression reduced 23 LEBA items to 3. All selected items were from Factor 3 (Using phone and smartwatch in bed): phone use before bed, phone check at night, and smartwatch check at night. The LASSO regression coefficients are presented in Table 2. Standard errors are not included as they are not meaningful for biased estimates arising from penalised estimation methods (Goeman et al., 2025). Variables shown in Table 2 were used to train the feature-reduced model.

**Table 2:** Least-absolute shrinkage and selection operator (LASSO) regression coefficients.

| Variable | Standardized Coefficients |
| --- | --- |
| Intercept | -0.18 |
| Phone use before bed | 0.31 |
| Phone check at night | 0.55 |
| Smartwatch check at night | 0.27 |

### Model Selection and Accuracy

The feature-reduced model (3 items) provided the highest cross-validation performance (mean AUC, 0.70 (SD = 0.13); mean sensitivity, 0.79 (SD = 0.18); mean specificity, 0.71 (SD = 0.23)) compared with the Factor 3 model using all 5 items (mean AUC, 0.68 (SD = 0.14); mean sensitivity, 0.73 (SD = 0.18); mean specificity, 0.78 (SD = 0.2)) and the full-feature model using all 23 LEBA items (mean AUC, 0.63 (SD = 0.12); mean sensitivity, 0.82 (SD = 0.18); mean specificity, 0.65 (SD = 0.22)).

When evaluated on the independent held-out test set (N = 23), the feature-reduced model achieved the highest AUC (0.83; sensitivity, 0.70; specificity, 0.92), followed by the Factor 3 model (AUC, 0.82; sensitivity, 0.70; specificity, 0.92) and the full-feature model (AUC, 0.79; sensitivity, 0.90; specificity, 0.77) (Table 3).

**Table 3:**
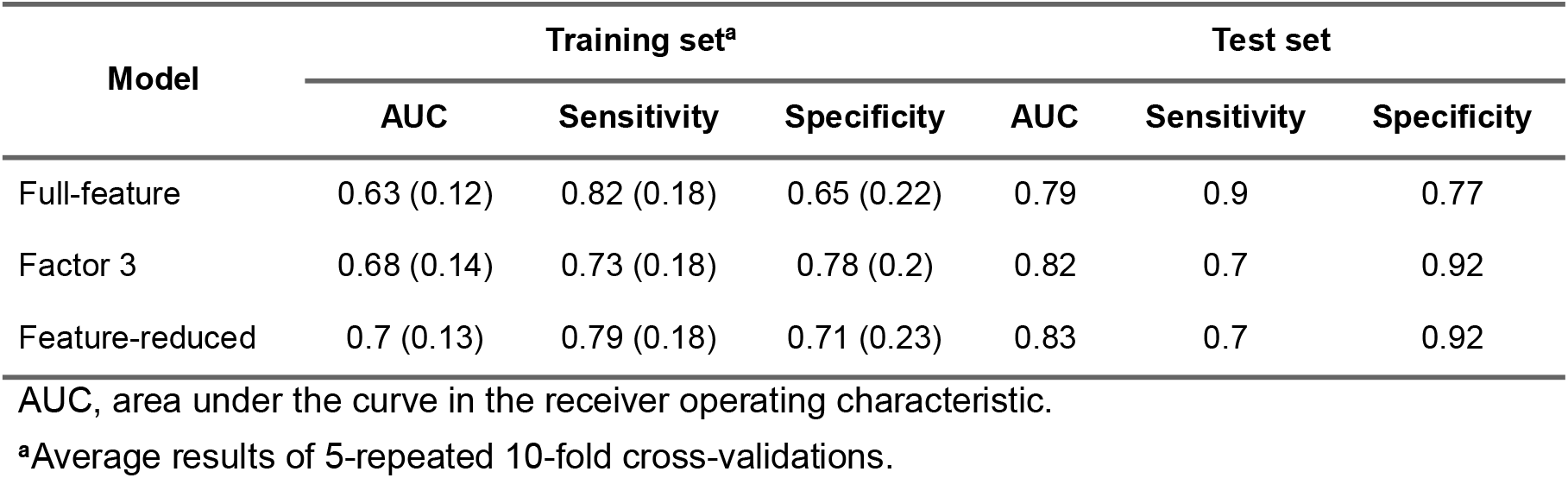
Comparison of performance models for training and test set.

| Model | Training set<sup>a</sup> |  |  | Test set |  |  |
| --- | --- | --- | --- | --- | --- | --- |
|  | AUC | Sensitivity | Specificity | AUC | Sensitivity | Specificity |
| Full-feature | 0.63 (0.12) | 0.82 (0.18) | 0.65 (0.22) | 0.79 | 0.9 | 0.77 |
| Factor 3 | 0.68 (0.14) | 0.73 (0.18) | 0.78 (0.2) | 0.82 | 0.7 | 0.92 |
| Feature-reduced | 0.7 (0.13) | 0.79 (0.18) | 0.71 (0.23) | 0.83 | 0.7 | 0.92 |

AUC, area under the curve in the receiver operating characteristic.
<sup>a</sup>Average results of 5-repeated 10-fold cross-validations.

### Nomogram

The nomogram was developed based on the feature-reduced model, as it achieved the highest predictive accuracy on the independent test set (Figure 2, Panel A). To support practical interpretation, a case example is included within the nomogram (Figure 2, Panel B).

**Figure 2:**
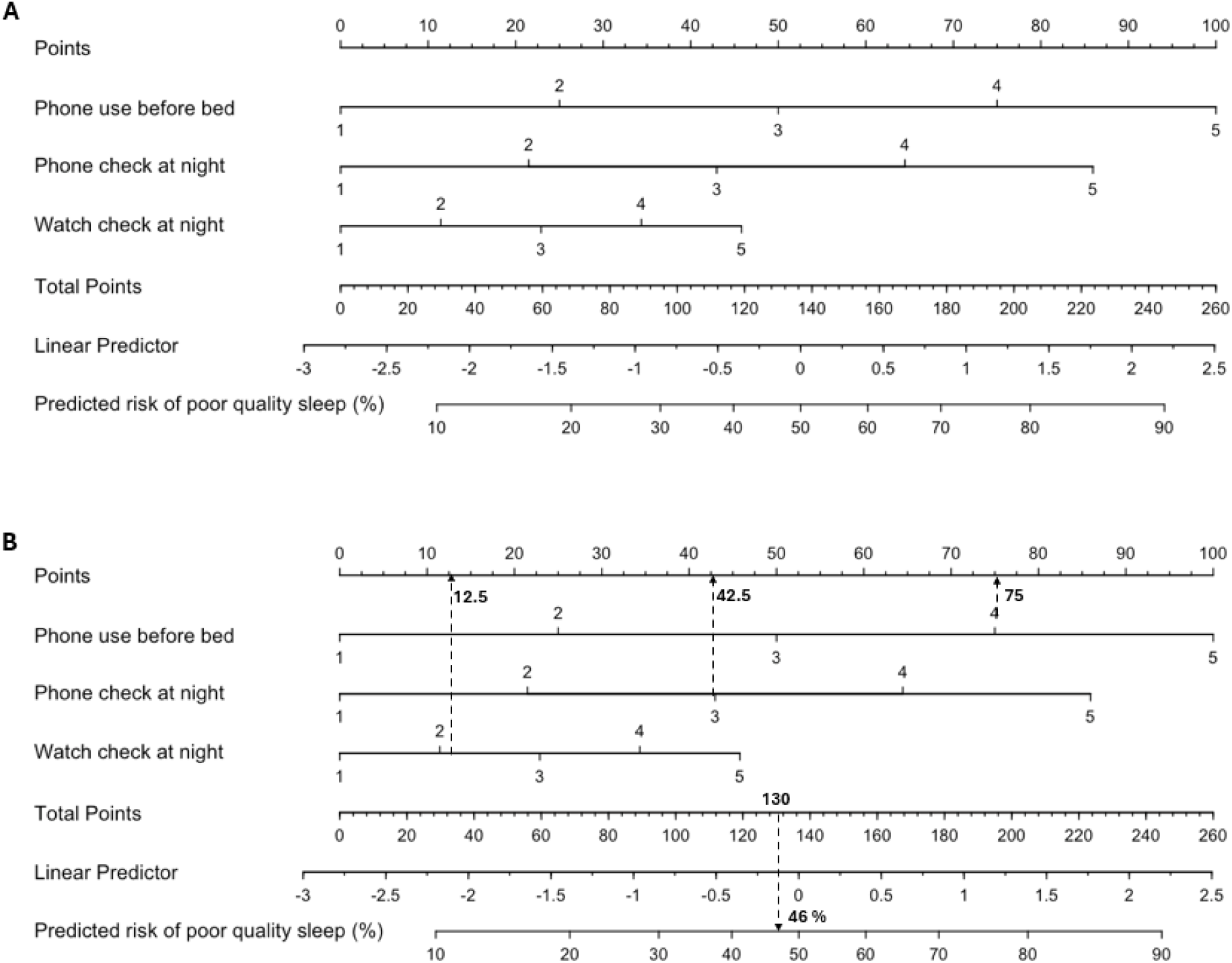
Nomogram for calculating the predicted risk of having poor sleep quality. Each predictor: phone use before bed, phone check at night, and watch check at night, is assigned a score based on its relative contribution to the model. Scores are summed to form the total points value, which is mapped to the linear predictor and the corresponding predicted chance of poor sleep quality. The Linear Predictor row shows the log-odds; most users can ignore this row and read directly from Total Points to Predicted Risk. **(A)** No case study annotations. **(B)** Case study scenario of an athlete who “often” (equivalent of 4 points on the LEBA’s 5-point Likert scoring scale) uses their mobile phone within 1 hour before attempting to fall asleep, “sometimes” (equivalent of 3 points) checks their phone when they wake up at night, and “rarely” (equivalent of 2 points) checks their smartwatch when they wake up at night. Extrapolating this information upward to the points obtained for each of these variables, such an individual would receive a total number of points of approximately 130, indicating a predicted risk of approximately 46 % of experiencing poor sleep quality, as determined by the Pittsburgh Sleep Quality Index (PSQI).

**How To Use This Nomogram:**

1. **Locate each predictor on its corresponding axis** Identify the individual’s response and assign scores based on the instrument’s 5-point Likert scoring scale (1 = never, 2 = rarely, 3 = sometimes, 4 = often, and 5 = always):
  - *Phone use before bed*
  - *Phone check at night*
  - *Smartwatch check at night*
2. **Assign points for each predictor** For each response category, draw a vertical line upward to the *Points* scale at the top of the nomogram and record the corresponding number of points.
3. **Calculate total points** Sum the points from all three predictors to obtain the *Total Points* value.
4. **Estimate risk of poor sleep quality** Locate the *Total Points* value on the corresponding axis and draw a vertical line downward to the *Predicted risk of poor quality sleep* scale to determine the estimated chance of poor sleep quality.

**Example (Panel B):**

An athlete who “often” uses their phone before bed (~75 points), “sometimes” checks their phone during the night (~42.5 points), and “rarely” checks their smartwatch at night (~12.5 points) would accumulate approximately **130 total points**, corresponding to an estimated **46% chance of poor sleep quality**. Therefore, the athlete in the current example has a 46% greater chance of obtaining a PSQI global score >5, indicating poor sleep quality.

## Discussion

This study sought to identify which light exposure behaviours were best associated PSQI-derived poor sleep quality and, from doing so, develop a practitioner-friendly visual tool to support light exposure behaviour evaluation in high-performance environments. Overall, our findings indicate that among the 23 Light Exposure Behaviour Assessment (LEBA) variables examined, the LASSO regression isolated three key predictors of sleep quality in athletes. These included: phone use before bed, phone check at night, and smartwatch check at night. These variables were subsequently incorporated into a nomogram designed to facilitate the practical interpretation of their combined association on sleep quality. Together, the results highlight that light-related behaviours may assist practitioners in identifying athletes at greater risk of poor sleep quality using minimal assessment time, which has practical implications for high-performance environments, particularly within routine athlete monitoring and screening.

### Phone use before bed

These findings align with evidence linking evening and nocturnal light exposure with poorer sleep outcomes and add nuance by identifying which behaviours are most relevant within athletic cohorts. Our findings report that using a phone before bed is associated with a higher likelihood of having poor sleep quality, which is partially consistent with the current literature. In general populations completing the LEBA, evening mobile phone use has been linked with circadian phase delays, reduced sleep quality, and cognitive difficulties (Siraji, Spitschan, et al., 2023b). In athletes, increased evening light exposure has likewise been associated with heightened sleep disturbances (Stevenson et al., 2024). However, in that study, the intensity of evening light exceeded light thresholds recommended by Brown et al., (2022), and the specific sources of exposure were not quantified. Systematic reviews report negative effects of blue-light exposure on sleep quality and total sleep time (Silvani et al., 2022). Although findings across the literature remain inconsistent. Recent consensus suggests that screen content, timing, engagement and individual light sensitivity may be more influential than spectral composition alone (Chellappa, 2021; Hartstein et al., 2024; Vézina-Im et al., 2025). Some findings further challenge assumptions about device use, for instance Driller & Uiga, (2019) reported no differences in subjective sleep quality between reading on an iPad versus a printed book before bedtime. Similarly, restricting evening device use combined with morning bright light did not significantly alter sleepiness in elite athletes (Hoshikawa, 2025). Accordingly, our findings may not be solely attributable to light emitted from smartphones but also the cognitively or emotionally stimulating content consumed before bed, which is increasingly recognised as a factor of sleep disruption (Levenson et al., 2016; Reichenberger et al., 2026).

### Phone/Smartwatch check at night

Results in the current study indicated that nighttime phone and smartwatch checking was also significantly associated with poor sleep quality in athletes. However, this association may be bidirectional, with device checking potentially occurring in response to nocturnal awakenings. Although photopic light exposure may contribute, behavioural and psychological mechanisms may be potentially involved. Notably, smartwatch checking may differ substantially from conventional watch checking in terms of light exposure, suggesting that the observed association may be driven by the act of checking itself rather than the light emission alone. For instance, frequent nighttime checking has been associated with poorer sleep and psychological wellbeing (Shoval et al., 2020). Further, “orthosomnia”, a phenomenon that is characterised by excessive endeavours to achieve optimal sleep metrics based on sleep statistics provided by sleep wearables, has become a recent topic highlighted in athletes (Driller et al., 2023; Jahrami et al., 2023; Trabelsi et al., 2023). Consensus statements warn that athletes may become overly focused on sleep or sleep-tracking data, potentially increasing sleep-related anxiety and checking behaviours (Walsh et al., 2021b). Nighttime checking behaviours may therefore reflect beyond the photic effects of brief light exposure, but potentially brief time checking or other screen activities e.g., scrolling on social media. However, the LEBA instrument does not capture the context in which these behaviours occur, and the cross-sectional design of the present study limits interpretation of the pathways underlying these associations. Accordingly, athletes experiencing sleep difficulties may engage in device checking because they are already awake and struggling to return to sleep, rather than device use causing their awakenings. The nomogram should therefore be interpreted as a screening tool to identify athletes exhibiting a behavioural profile associated with poorer sleep quality rather than a diagnostic tool or guide for specific light-based interventions. Therefore, longitudinal and multi-timepoint monitoring may help clarify this relationship and determine whether these behaviours are a cause or consequence of sleep disruption. Additionally, these behaviours may also reflect awakenings driven by training stress, scheduling demands or other non-light-related factors. Athlete sleep behaviours vary substantially with training load, scheduling, and competition demands (Nedelec et al., 2018), and light exposure patterns may also differ by sport. For instance, Stevenson et al., (2024) reported markedly higher outdoor light exposure in team-sport athletes, levels of which are noticeably exceeding those observed in general populations (Didikoglu et al., 2023). Such exposure may buffer the effects of suboptimal evening light behaviours (Brown et al., 2022). Future work should examine these interactions by considering overall 24-hour light exposure, sport-specific contexts, and the factors influencing nocturnal awakenings.

### Comparison of classification models

The feature-reduced model produced the highest predictive accuracy compared to the Factor 3 and full-feature models. This likely reflects reduced model complexity and decreased risk of overfitting (McNeish, 2015), as increasing the number of predictors increases the likelihood of capturing noise rather than meaningful variance. In practical terms, a model with fewer, more informative predictors can generalise more reliably to new athletes because it has learned the underlying pattern rather than memorising idiosyncrasies of the training sample. Feature reduction can therefore be considered to refine athlete monitoring practices by reducing questionnaire burden while retaining predictive utility. Rather than administering the full LEBA questionnaire, practitioners may need only assess three behaviours: phone use before bed, phone checking at night, and smartwatch checking at night, to screen for athletes who may be at risk of poor sleep quality. Practitioners may consider the feature-reduced model to optimise monitoring and to focus feedback on the most meaningful light-related behaviours.

### Nomogram

The second aim of the present study was to develop a practitioner-friendly nomogram to translate statistically significant light-exposure predictors into an accessible visual tool of sleep quality. By integrating significant predictors, this nomogram allows applied practitioners to screen for behavioural profiles associated with poor sleep quality in athletes. The proposed nomogram encompasses items from Factor 3 of the LEBA tool, which is specifically relevant to in-bed screen-related and nighttime phone and smartwatch checking. Given the increasing emphasis on monitoring wellness, sleep and performance in elite sport (Halson, 2019), this nomogram provides a convenient, low-burden tool to screen athletes at elevated risk of poor sleep and support regular light-related assessments. This tool’s ease of use and initial predictive accuracy support its value in applied sport settings. To the author’s knowledge, no existing athlete-focused efficient screening tool provides a visual, behaviour-based predictive model of sleep quality, highlighting the potential value of the present nomogram as a complementary tool within athlete monitoring systems. Practitioners could administer the three LEBA items (phone use before bed, phone check at night, smartwatch check at night) as a brief screening tool during athlete consultations. Higher nomogram scores should prompt practitioners to investigate why the athlete is experiencing sleep difficulties, with device checking viewed as a marker rather than necessarily a primary intervention target. Future research should evaluate the implementation of this nomogram in high-performance settings and examine its utility as part of athlete sleep education initiatives targeting light exposure behaviours.

### Limitations and Future Research

There are several limitations that should be considered when interpreting the results of this study. First, the subjective nature of the tools used introduces potential recall and response bias. Athletes were asked to recall behaviours “over the past four weeks”, which athletes may have difficulty precisely recalling both light exposure-related and sleep-related behaviours over that time. Further, global distribution of respondents in this study, light exposure, particularly outdoor light intensity, may have varied substantially depending on climate, hemisphere (northern vs. southern), and time of year (e.g., summer vs. winter). Although the LASSO regression identified predominantly indoor light behaviours as most strongly associated with poor sleep quality, seasonal and geographical differences in light exposure may still influence overall light habits and sleep outcomes. Therefore, future research incorporating more detailed information on climate and season with standardised follow-up assessments across different times of year is warranted to evaluate the generalisability and predictive accuracy of the developed nomogram. Additionally, only subjective measures of light exposure and sleep were collected, and sleep quality was defined via the PSQI. Athlete-specific questionnaires (e.g., Athlete Sleep Screening Questionnaire or Athlete Sleep Behaviour Questionnaire) may have yielded different insights. Further, the inclusion of objective tools (e.g., wearable light sensors) would enable quantification of light intensity, spectral composition, and temporal patterns. Future research should examine the validity of the nomogram against such measures to gain further insight into the mechanisms underpinning these light-related behaviours. Finally, the cross-sectional design of the current study limits causal inference. Future research should examine targeted light-related interventions based on the feature-reduced model on sleep outcomes in athletes and incorporate salivary melatonin measurements to clarify underlying mechanisms. In addition, future research should compare nomograms and predictive variables across different athlete populations (e.g., elite vs. highly trained) to determine whether light exposure behaviours generalise across performance levels. Future work should also examine whether the identified variables better predict broader sleep and circadian outcomes, such as sleep regularity and physiological markers, to provide a more comprehensive understanding of how light exposure behaviours influence circadian rhythms, beyond subjective sleep quality alone.

## Conclusion

This study identified three key light-related behaviours that are most strongly associated with subjective sleep quality in athletes and demonstrated that feature reduction can clarify multivariate influences. Athletes who frequently use phones before bed, check phones during night awakenings, and check smartwatches at night are at elevated risk of poor sleep quality. These three behaviours, captured by a brief screening, provide practitioners with actionable targets for light-related sleep assessments. Athlete self-report tools remain cost-effective and practical, but can be burdensome when numerous variables are collected. Therefore, the proposed nomogram provides a convenient and low-burden approach to identifying athletes who may be susceptible to poor sleep quality due to their light exposure habits. Further, identifying these key behaviours and the nomogram provides a foundation for screening athletes at risk of poor sleep quality, supporting practitioner efforts to refine education and optimise athlete sleep and circadian health.

## Data Availability

All data produced in the present study are available upon reasonable request to the authors

## References

1. Didikoglu, N. Mohammadian, S. Johnson, M. van Tongeren, P. Wright, A. Casson, T. Brown, and R. L. (2023). Associations between light exposure and sleep timing and sleepiness while awake in a sample of UK adults in everyday life. Proceedings of the National Academy of Sciences, 120(42), 1–9. 10.1073/pnas

2. Bajaj, A., Rosner, B., Lockley, S. W., & Schernhammer, E. S. (2011). Validation of a light questionnaire with real-life photopic illuminance measurements: The harvard light exposure assessment questionnaire. Cancer Epidemiology Biomarkers and Prevention, 20(7), 1341–1349. 10.1158/1055-9965.EPI-11-0204

3. Brown, T. M., Brainard, G. C., Cajochen, C., Czeisler, C. A., Hanifin, J. P., Lockley, S. W., Lucas, R. J., Münch, M., OHagan, J. B., Peirson, S. N., Price, L. L. A., Roenneberg, T., Schlangen, L. J. M., Skene, D. J., Spitschan, M., Vetter, C., Zee, P. C., & Wright, K. P. (2022). Recommendations for daytime, evening, and nighttime indoor light exposure to best support physiology, sleep, and wakefulness in healthy adults. PLoS Biology, 20(3). 10.1371/journal.pbio.3001571

4. Burns, A. C., Saxena, R., Vetter, C., Phillips, A. J. K., Lane, J. M., & Cain, S. W. (2021). Time spent in outdoor light is associated with mood, sleep, and circadian rhythm-related outcomes: A cross-sectional and longitudinal study in over 400,000 UK Biobank participants. Journal of Affective Disorders, 295(June), 347–352. 10.1016/j.jad.2021.08.056

5. Buysse, D. J., Reynolds, C. F., Monk, T. H., Berman, S. R., & Kupfer, D. J. (1989). The Pittsburgh Sleep Quality Index: a new instrument for psychiatric practice and research.

6. Chellappa, S. L. (2021). Individual differences in light sensitivity affect sleep and circadian rhythms. In Sleep (Vol. 44, Issue 2). Oxford University Press. 10.1093/sleep/zsaa214

7. Corbett, R. W., Middleton, B., & Arendt, J. (2012). An hour of bright white light in the early morning improves performance and advances sleep and circadian phase during the Antarctic winter. Neuroscience Letters, 525(2), 146–151. 10.1016/j.neulet.2012.06.046

8. Driller, M., & Uiga, L. (2019). The influence of night-time electronic device use on subsequent sleep and propensity to be physically active the following day. Chronobiology International, 36(5), 717–724. 10.1080/07420528.2019.1588287

9. Duffy, J. F., & Czeisler, C. A. (2009). Effect of Light on Human Circadian Physiology. In Sleep Medicine Clinics (Vol. 4, Issue 2, pp. 165–177). 10.1016/j.jsmc.2009.01.004

10. Duffy, J. F., & Wright, K. P. (2005). Entrainment of the human circadian system by light. In Journal of Biological Rhythms (Vol. 20, Issue 4, pp. 326–338). 10.1177/0748730405277983

11. Figueiro, M. G., Steverson, B., Heerwagen, J., Kampschroer, K., Hunter, C. M., Gonzales, K., Plitnick, B., & Rea, M. S. (2017). The impact of daytime light exposures on sleep and mood in office workers. Sleep Health, 3(3), 204–215. 10.1016/j.sleh.2017.03.005

12. Friedman, J., Hastie, T., & Tibshirani, R. (2010). Regularization Paths for Generalized Linear Models via Coordinate Descent. In JSS Journal of Statistical Software (Vol. 33). http://www.jstatsoft.org/

13. Goeman, J., Meijer, R., & Chaturvedi, N. (2025). L1 and L2 Penalized Regression Models.

14. Halson, S. L. (2019). Sleep Monitoring in Athletes: Motivation, Methods, Miscalculations and Why it Matters. Sports Medicine, 49(10), 1487–1497. 10.1007/s40279-019-01119-4

15. Harrell Jr, F. E. (2023). rms: Regression Modeling Strategies. R package version 6. 2-0. 2021.

16. Hartstein, L. E., Mathew, G. M., Reichenberger, D. A., Rodriguez, I., Allen, N., Chang, A. M., Chaput, J. P., Christakis, D. A., Garrison, M., Gooley, J. J., Koos, J. A., Van Den Bulck, J., Woods, H., Zeitzer, J. M., Dzierzewski, J. M., & Hale, L. (2024). The impact of screen use on sleep health across the lifespan: A National Sleep Foundation consensus statement. In Sleep Health (Vol. 10, Issue 4, pp. 373–384). Elsevier Inc. 10.1016/j.sleh.2024.05.001

17. Hastings, M. H., Maywood, E. S., & Reddy, A. B. (2008). Two decades of circadian time. Journal of Neuroendocrinology, 20(6), 812–819. 10.1111/j.1365-2826.2008.01715.x

18. Hoshikawa, M. (2025). Effects of Nightly Electronic Device Restriction and Morning Bright Light on Sleep, Mood, and Performance Among Elite Athletes. https://www.nsca.com

19. Jones, M. J., Dawson, B., Gucciardi, D. F., Eastwood, P. R., Miller, J., Halson, S. L., Dunican, I. C., & Peeling, P. (2019). Evening electronic device use and sleep patterns in athletes. Journal of Sports Sciences, 37(8), 864–870. 10.1080/02640414.2018.1531499

20. Knufinke, M., Fittkau-Koch, L., Møst, E. I. S., Kompier, M. A. J., & Nieuwenhuys, A. (2019a). Restricting short-wavelength light in the evening to improve sleep in recreational athletes–A pilot study. European Journal of Sport Science, 19(6), 728–735. 10.1080/17461391.2018.1544278

21. Knufinke, M., Nieuwenhuys, A., Geurts, S. A. E., Møst, E. I. S., Moen, M. H., Maase, K., Coenen, A. M. L., Gordijn, M. C. M., & Kompier, M. A. J. (2021). Dim light, sleep tight, and wake up bright–Sleep optimization in athletes by means of light regulation. European Journal of Sport Science, 21(1), 7–15. 10.1080/17461391.2020.1722255

22. Kuhn, M. (2015). Caret: classification and regression training. Astrophysics Source Code Library, ascl-1505.

23. McKay, A. K. A., Stellingwerff, T., Smith, E. S., Martin, D. T., Mujika, I., Goosey-Tolfrey, V. L., Sheppard, J., & Burke, L. M. (2022). Defining Training and Performance Caliber: A Participant Classification Framework. International Journal of Sports Physiology and Performance, 17(2), 317–331. 10.1123/ijspp.2021-0451

24. McNeish, D. M. (2015). Using Lasso for Predictor Selection and to Assuage Overfitting: A Method Long Overlooked in Behavioral Sciences. Multivariate Behavioral Research, 50(5), 471–484. 10.1080/00273171.2015.1036965

25. Nedelec, M., Aloulou, A., Duforez, F., Meyer, T., & Dupont, G. (2018). The Variability of Sleep Among Elite Athletes. In Sports Medicine - Open (Vol. 4, Issue 1). Springer. 10.1186/s40798-018-0151-2

26. O’Donnell, S., & Driller, M. W. (2017). Sleep-hygiene Education improves Sleep Indices in Elite Female Athletes. International Journal of Exercise Science, 10(4), 522–530.

27. Sargent, C., Lastella, M., Halson, S. L., & Roach, G. D. (2014). The impact of training schedules on the sleep and fatigue of elite athletes. Chronobiology International, 31(10), 1160–1168. 10.3109/07420528.2014.957306

28. Scheuermaier, K., Laffan, A. M., & Duffy, J. F. (2010). Light exposure patterns in healthy older and young adults. Journal of Biological Rhythms, 25(2), 113–122. 10.1177/0748730410361916

29. Shoval, D., Tal, N., & Tzischinsky, O. (2020). Relationship of smartphone use at night with sleep quality and psychological well-being among healthy students: A pilot study. Sleep Health, 6(4), 495–497. 10.1016/j.sleh.2020.01.011

30. Silvani, M. I., Werder, R., & Perret, C. (2022). The influence of blue light on sleep, performance and wellbeing in young adults: A systematic review. In Frontiers in Physiology (Vol. 13). Frontiers Media S.A. 10.3389/fphys.2022.943108

31. Siraji, M. A., Lazar, R., Duijnhoven, J., Schlangen, L., Haque, S., Kalavally, V., Vetter, C., Glickman, G., Smolders, K., & Spitschan, M. (2022a). Light exposure behaviour assessment (LEBA): A novel self-reported instrument to capture light exposure-related behaviour.

32. Siraji, M. A., Lazar, R. R., van Duijnhoven, J., Schlangen, L. J. M., Haque, S., Kalavally, V., Vetter, C., Glickman, G. L., Smolders, K. C. H. J., & Spitschan, M. (2023). An inventory of human light exposure behaviour. Scientific Reports, 13(1), 1–15. 10.1038/s41598-023-48241-y

33. Siraji, M. A., Spitschan, M., Kalavally, V., & Haque, S. (2023a). Light exposure behaviors predict mood, memory and sleep quality. Scientific Reports, 13(1), 1–14. 10.1038/s41598-023-39636-y

34. Stevenson, S., Suppiah, H., & Driller, M. (2025). Research note: The validity and reliability of a novel wearable sensor in measuring light exposure in an indoor and outdoor environment. Lighting Research and Technology. 10.1177/14771535251338582

35. Stevenson, S., Suppiah, H., Ruddy, J., Murphy, S., & Driller, M. (2024). Higher Levels of Morning and Daytime Light Exposure Associated with Positive Sleep Indices in Professional Team Sport Athletes. August, 1279–1290.

36. Stone, J. E., McGlashan, E. M., Facer-Childs, E. R., Cain, S. W., & Phillips, A. J. K. (2020). Accuracy of the GENEActiv Device for Measuring Light Exposure in Sleep and Circadian Research. Clocks & Sleep, 2(2), 143–152. 10.3390/clockssleep2020012

37. Suppiah, H. T., Swinbourne, R., Wee, J., He, Q., Pion, J., Driller, M. W., Gastin, P. B., & Carey, D. L. (2022). Predicting Youth Athlete Sleep Quality and the Development of a Translational Tool to Inform Practitioner Decision Making. Sports Health, 14(1), 77–83. 10.1177/19417381211056078

38. Vézina-Im, L. A., Morin, C. M., Chen, S., Ivers, H., Carney, C. E., Chaput, J. P., Dang-Vu, T. T., Davidson, J. R., & Robillard, R. (2025). The complex association between bedtime screen use and adult sleep health. Sleep Health, 11(5), 572–578. 10.1016/j.sleh.2025.06.010

39. Vitale, J. A., Banfi, G., Sias, M., & La Torre, A. (2019). Athletes’ rest-activity circadian rhythm differs in accordance with the sport discipline. Chronobiology International, 36(4), 578–586. 10.1080/07420528.2019.1569673

40. Walsh, N. P., Halson, S. L., Sargent, C., Roach, G. D., Nédélec, M., Gupta, L., Leeder, J., Fullagar, H. H., Coutts, A. J., Edwards, B. J., Pullinger, S. A., Robertson, C. M., Burniston, J. G., Lastella, M., Le Meur, Y., Hausswirth, C., Bender, A. M., Grandner, M. A., & Samuels, C. H. (2021a). Sleep and the athlete: Narrative review and 2021 expert consensus recommendations. British Journal of Sports Medicine, 55(7), 356–368. 10.1136/bjsports-2020-102025

41. Watson, A. M. (2017). Sleep and Athletic Performance. Current Sports Medicine Reports, 16(6), 413–418. 10.1249/JSR.0000000000000418

42. Yin, J., Julius, A. A., & Wen, J. T. (2021). Optimization of light exposure and sleep schedule for circadian rhythm entrainment. PLoS ONE, 16(6 June). 10.1371/journal.pone.0251478

